# Outcomes of Hydroxychloroquine Treatment Among Hospitalized COVID-19 Patients in the United States- Real-World Evidence From a Federated Electronic Medical Record Network

**DOI:** 10.1101/2020.05.12.20099028

**Authors:** Shailendra Singh, Ahmad Khan, Monica Chowdhry, Arka Chatterjee

## Abstract

On March 28, 2020, in response to the rapidly accelerating COVID-19 pandemic, U.S FDA issued emergency use authorization for hydroxychloroquine (HCQ) in hospitalized COVID-19 patients based on limited in-vitro and anecdotal clinical data. Analysis of the accumulated real-world data utilizing electronic medical records (EMR) could indicate HCQ therapy benefits as we await the results of clinical trials. However, any such analysis of retrospective observational data should account for variables such as demographics and comorbidities that could affect treatment strategies or outcomes. Therefore, we report the outcomes of HCQ treatment in a propensity-matched cohort of COVID-19 hospitalized patients. Our analysis of a large retrospective cohort of hospitalized COVID-19 patients treated with HCQ did not show benefits in mortality or the need for mechanical ventilation when compared to a matched cohort of patients who did not receive HCQ.

## Introduction

On March 28, *2020*, in response to the rapidly accelerating COVID-19 pandemic, U.S FDA issued emergency use authorization for hydroxychloroquine (HCQ) in hospitalized COVID-19 patients based on limited in-vitro and anecdotal clinical data^1,2^. Analysis of the accumulated real-world data utilizing electronic medical records (EMR) could indicate HCQ therapy benefits as we await the results of clinical trials. However, any such analysis of retrospective observational data should account for variables such as demographics and comorbidities that could affect treatment strategies or outcomes. Therefore, we report the outcomes of HCQ treatment in a propensity-matched cohort of COVID-19 hospitalized patients.

## Methods and Findings

Using the TriNetX (Cambridge, MA, USA), a global federated health research network, we performed a real-time search and analysis of EMR of more than 40 million patients from 34 healthcare organizations (HCOs) in the United States. TriNETX recently fast-tracked data inflow to incorporate COVID-19 specific diagnosis and terminology following the World Health Organization (WHO) and Centers for Disease Control (CDC) criteria. As a federated network, TriNetX received a waiver from Western IRB since only aggregated counts, statistical summaries of de-identified information, but no protected health information is received, and no study-specific activities are performed in retrospective analyses.

All hospitalized adult patients (≥ 18 years) diagnosed with COVID-19 between January 20, *2020*, and May 1, 2020, were identified using COVID-19 specific diagnosis and laboratory findings following the WHO and CDC COVID-19 guidelines (N=3618). We excluded patients who received potential COVID-19 specific therapeutic agents (N=246) other than HCQ (Azithromycin and corticosteroids were not considered COVID-19 specific) (Figure 1). Remaining patients (N=3372) were then stratified into two groups: HCQ (Treatment group; N=1125) and non-HCQ group (control group; N=2247) (Figure 1). Details of data source, coding systems to present data, definition of variables and statistical analysis can be found in the Supplementary Methods.

**Figure 1:**
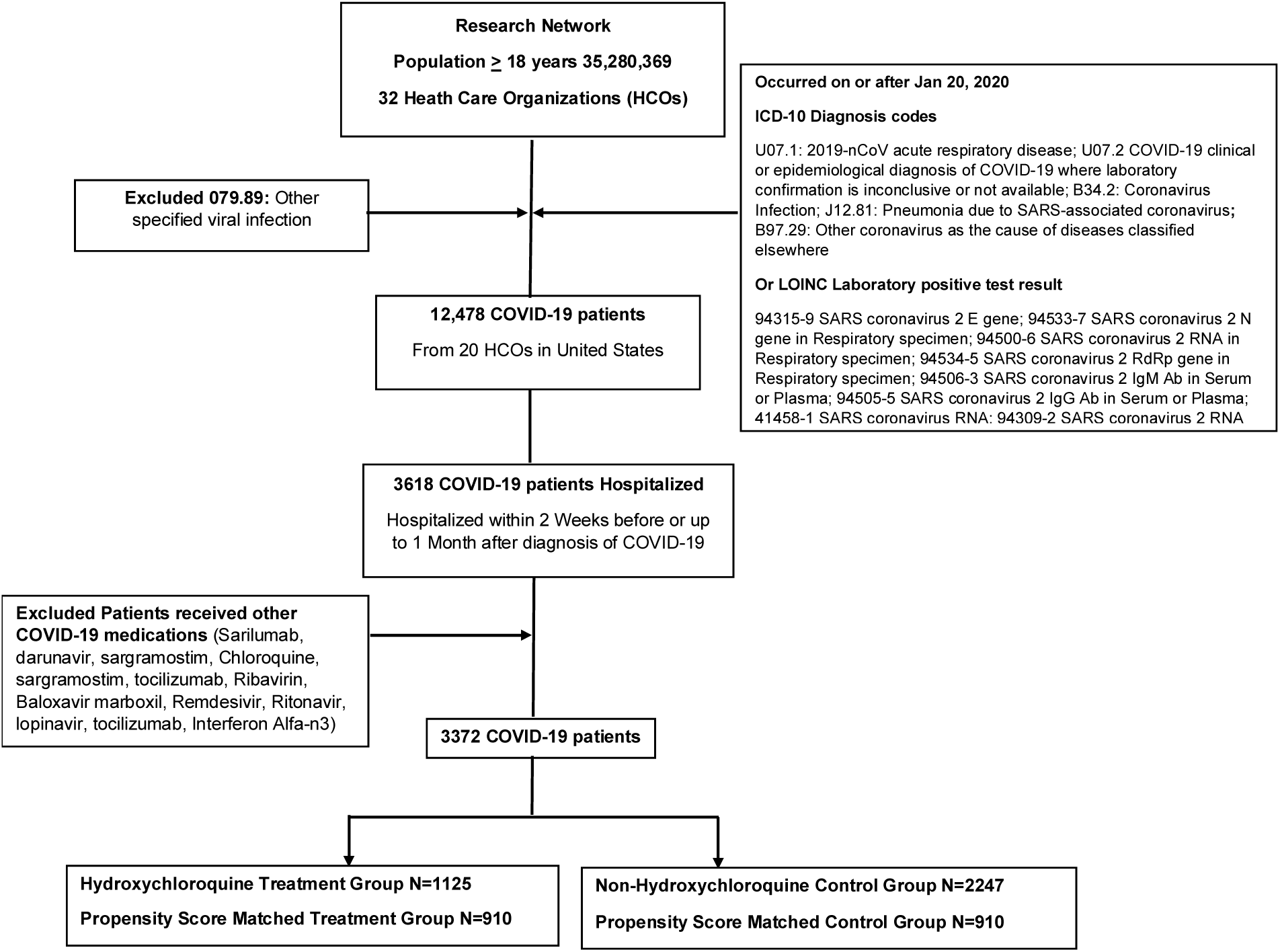
Flow chart of patient selection in the Hydroxychloroquine treatment group and Control group.

Because of significant differences in demographics and comorbidities between the treatment and control groups, we performed propensity score matching for age, gender, race, and potential confounding comorbidities. After propensity matching, relatively balanced cohorts of 910 patients in each group were selected for further analysis (Table 1) (Supplementary Figure 1). Mortality and need for mechanical ventilation in the treatment and control groups were similar (Table 1). We performed a sensitivity analysis to test the robustness of these findings. The estimated risk of 7-day (RR 1.00, 0.73-1.37), and 14-day (RR 1.04, 95% CI 0.80-1.36) mortality after the start of HCQ treatment were similar. The mortality was also not significantly different when cohorts were only matched for age, gender, race, diabetes, and hypertension (N=982) (RR 0.93, 95%CI 0.78-1.19). HCQ was used in combination with Azithromycin in a majority of patients (N=799). When we compared a matched cohort of HCQ and Azithromycin combination with the control group (N=701), we did not see any benefit in mortality, or mechanical ventilation. Incidence of new events of ventricular tachycardia/ fibrillation or sudden cardiac death was seen in 1.09% (N=10) patients in the HCQ treatment, which was similar to the control group (RR 0.63, 95%CI 0.28-1.37).

**Table 1:** Comparison of patient characteristics and outcomes among hospitalized COVID-19 Hydroxychloroquine Treatment group and Control group. Patient demographics and baseline comorbidities are compared before and after propensity matching of groups.

| Demographics and Comorbidities |  |  |  |  |  |  |
| --- | --- | --- | --- | --- | --- | --- |
|  | Before propensity matching |  |  | After propensity matching |  |  |
|  | Treatment Group<br>(n=1125) | Control Group<br>(n=2247) | P-value | Treatment Group<br>(n=910) | Control Group<br>(n=910) | P-value |
| Mean Age years (S.D) | 61.45 ±16.60 | 62.30 ±17.04 | 0.17 | 62.17±16.81 | 62.55±17.62 | 0.63 |
| Male % (N) | 56.09% (631) | 49.71% (1117) | <0.001 | 53.96% (491) | 54.94% (500) | 0.67 |
| Race |  |  |  |  |  |  |
| White % (N) | 51.20% (576) | 38.41% (863) | <0.001 | 57.25% (521) | 58.68% (534) | 0.54 |
| Black or African American % (N) | 18.40% (207) | 52.87% (1188) | <0.001 | 22.64% (206) | 21.10% (192) | 0.43 |
| Unknown Race % (N) | 28.89% (325) | 7.48% (168) | <0.001 | 18.35% (167) | 18.02% (164) | 0.86 |
| Hypertensive diseases<br>% (N) | 61.42% (691) | 64.44% (1448) | 0.086 | 62.75% (571) | 60.33% (549) | 0.29 |
| Diabetes mellitus %<br>(N) | 37.07% (417) | 37.78% (849) | 0.68 | 36.92% (336) | 33.63% (306) | 0.14 |
| Obesity % (N) | 34.76% (391) | 20.69% (465) | <0.001 | 31.54% (287) | 28.90% (263) | 0.22 |
| Ischemic heart<br>diseases % (N) | 28.89% (325) | 24.48% (550) | 0.006 | 28.79% (262) | 28.90% (263) | 0.96 |
| Chronic kidney<br>disease (CKD) % (N) | 21.16% (238) | 27.33% (614) | <0.001 | 23.41% (213) | 21.43% (195) | 0.31 |
| Heart failure % (N) | 16.71% (188) | 22.65% (509) | <0.001 | 19.12% (174) | 18.13% (165) | 0.59 |
| Prolonged QT interval<br>or % Long QT<br>Syndrome (N) | 2.22% (25) | 2.58% (58) | 0.53 | 2.31% (21) | 2.75% (25) | 0.55 |
| Atrial fibrillation and flutter % (N) | 14.93% (168) | 17.05% (383) | 0.12 | 16.59% (151) | 17.25% (157) | 0.71 |
| Cerebrovascular diseases % (N) | 12.98% (146) | 17.13% (385) | 0.002 | 14.94% (136) | 14.94% (136) | 1.00 |
| Chronic obstructive pulmonary disease % (N) | 13.16% (148) | 14.15% (318) | 0.43 | 14.50% (132) | 13.96% (127) | 0.74 |
| Asthma % (N) | 12.89% (145) | 12.33% (277) | 0.64 | 13.30% (121) | 12.97% (118) | 0.84 |
| Diseases of liver % (N) | 8.89% (100) | 12.32% (277) | 0.003 | 10.22% (93) | 9.67% (88) | 0.70 |
| Malignant neoplasms of lymphoid, hematopoietic and related tissue % (N) | 3.20% (36) | 3.29% (74) | 0.89 | 3.19% (29) | 3.63% (33) | 0.61 |
| Rheumatoid arthritis % (N) | 3.11% (35) | 1.78% (40) | 0.013 | 2.75% (25) | 2.64% (24) | 0.88 |
| Malignant neoplasm<br>of breast % (N) | 2.22% (25) | 2.00% (45) | 0.67 | 2.53% (23) | 2.31% (21) | 0.76 |
| Malignant neoplasm<br>of colon % (N) | 1.78% (20) | 1.16% (26) | 0.14 | 1.87% (17) | 1.65% (15) | 0.72 |
| Malignant neoplasm<br>of prostate % (N) | 1.51% (17) | 2.40% (54) | 0.089 | 1.65% (15) | 1.76% (16) | 0.86 |
| Systemic lupus<br>erythematosus (SLE)<br>% (N) | 1.69% (19) | 0.89% (20) | 0.041 | 1.32% (12) | 1.43% (13) | 0.84 |
| Human<br>immunodeficiency<br>virus [HIV] disease %<br>(N) | 0.89% (10) | 0.80% (18) | 0.79 | 1.10% (10) | 1.10% (10) | 1.00 |
| Nicotine dependence<br>% (N) | 11.20% (126) | 11.08% (249) | 0.92 | 11.87% (108) | 10.99% (100) | 0.56 |
| Alcohol related disorders % (N) | 4.62% (52) | 4.36% (98) | 0.73 | 4.72% (43) | 4.72% (43) | 1.00 |

Laboratory Findings after COVID-19 Hospitalization in Matched Cohort
| Laboratory Finding<br>(Serum, Plasma or Blood) | Treatment Group<br>n=910<br>Mean ± S.D (N) | Control Group<br>n=910<br>Mean ± S.D (N) | P-Value |
| --- | --- | --- | --- |
| Alanine aminotransferase (ALT) (U/L) | 46.24 ± 60.73 (175) | 88.68 ± 366.75 (495) | 0.128 |
| Total Bilirubin (mg/dL) | 0.56 ± 0.42 (175) | 0.82 ± 1.19 (491) | 0.007 |
| Creatinine (mg/dL) | 1.54 ± 1.84 (270) | 1.61 ± 2.05 (578) | 0.60 |
| Leukocytes (/mcL) | 7.80 ± 4.74 (244) | 9.41 ± 6.40 (563) | 0.0004 |
| Lymphocytes (/mCL) | 1.30 ± 0.65<br>(195) | 1.78 ± 3.29 (150) | 0.0454 |
| Prothrombin time (PT)<br>(seconds) | 16.61 ± 7.25 (70) | 15.33 ± 10.08<br>(165) | 0.3389 |
| Ferritin (ng/mL) | 1119.74 ±<br>1656.77 (150) | 1693.40 ± 2656.00<br>(202) | 0.02 |
| C reactive protein<br>(mg/L) | 19.45 ± 41.11<br>(152) | 98.8 ± 101.37<br>(350) | <0.0001 |
| Erythrocyte<br>sedimentation rate<br>(mm/hr) | 56.53 ± 30.03 (17) | 57.93 ± 33.64 (57) | 0.88 |
| Lactate dehydrogenase<br>(U/L) | 373.80 ± 160.39<br>(145) | 475.21 ± 814.75<br>(155) | 0.14 |
| Interleukin-6 (pg/mL) | 70.00 ± 80.57 (18) | 223.73 ± 853.07<br>(27) | 0.45 |
| Procalcitonin (ng/mL) | 3.39 ± 11.41 (25) | 2.64 ± 8.30 (118) | 0.70 |

Treatment Hydroxychloroquine vs Control (Matched Cohorts)
| Outcomes | Treatment Group | Control group | Relative Risk | Risk Difference | P-Value |
| --- | --- | --- | --- | --- | --- |
|  | n=910<br>% (N) | n=910<br>% (N) | (95% CI) | (95% CI) |  |
| Mortality 30-Day | 11.43% (104) | 11.98% (109) | 0.95<br>(0.74,1.23) | -0.55% (-<br>3.50%,2.40%) | 0.72 |
| Mechanical Ventilation | 5.05% (46) | 6.26% (57) | 0.81<br>(0.55,1.18) | -1.21% (-<br>3.33%,0.91%) | 0.26 |

Treatment Hydroxychloroquine combined with Azithromycin vs. Control (Matched Cohorts)
| Outcomes | Treatment | Control group |  | Risk Difference | P-Value |
| --- | --- | --- | --- | --- | --- |
|  | n=701 | n=701 | Risk Ratio | (95% CI) |  |

|  | % (N) | % (N) | (95% CI) |  |  |
| --- | --- | --- | --- | --- | --- |
| Mortality | 12.27% (86) | 10.27% (72) | 1.19<br>(0.89,1.60) | 2.00% (-<br>1.31%,5.30%) | 0.24 |
| Mechanical ventilation | 5.71% (40) | 5.85% (41) | 0.976<br>(0.64,1.49) | -0.14% (-<br>2.58%,2.30%) | 0.91 |

n=Total number of patients, N=patients with outcomes

## Discussion

The interest in the therapeutic use of HCQ in COVID19 stems from the evidence of in-vitro antiviral activity against SARS-Coronavirus 2^1^. Results from a small non-randomized study showed a higher rate of viral load reduction after HCQ treatment^2^. However, the apparent antiviral activity of HCQ does not seem to have translated into clinical outcomes^3,4^. Our analysis of a large retrospective cohort of hospitalized COVID-19 patients treated with HCQ did not show benefits in mortality or the need for mechanical ventilation when compared to a matched cohort of patients who did not receive HCQ. Without proven efficacy, serious adverse events reported with HCQ use in COVID-19 is of foremost concern^5^. Ventricular Arrhythmic complications were reported in a small number of COVID-19 patients in our study. While real-world data drawn from EHR have limitations, our study still provides valuable evidence against the widespread use of HCQ, once described as a *“game-changer,”* without proven benefits in randomized controlled studies.

## Data Availability

Details of data source and code used are available in the supplementary

## Supplementary

### Methods

**Figure 1:** Propensity Score Density Graph before and after propensity score matching of the Hydroxychloroquine treatment group and the control group.

## Acknowledgment

We acknowledge the West Virginia Clinical and Translational Science Institute to provide us access, and training to the TriNETXglobal healthcare network.

We also acknowledge the TriNETX (Cambridge, MA, USA) healthcare network for design assistance to complete this project.

The authors received no financial support or grants for the research, authorship, and publication of this article.

Shailendra Singh, Ahmad Khan, Monica Chowdhry and Arka Chatterjee declare that they have no conflict of interest.

